# Dynamics of Drug Overdose Deaths in the United States During COVID-19

**DOI:** 10.1101/2025.05.17.25327834

**Authors:** Hawre Jalal, Donald S. Burke

## Abstract

**Importance:** Drug overdose (OD) deaths in the United States have risen exponentially for four decades and surged further during the COVID 19 pandemic; the drivers of this surge remain unclear.

**Objective:** To quantify excess OD mortality during COVID 19, separate its temporal components, and identify social, economic, and drug supply factors—especially Economic Impact Payments (EIPs)—linked to those components.

**Design, Setting, and Participants:** Temporal and spatial analysis of all 50 U.S. states using OD deaths (1979–2023) merged with state level data on COVID-19, unemployment, income, mobility, fentanyl seizures, opioid treatment supply, and EIP timing/amounts.

**Methods:** A log linear model projected the 40-year exponential trend and seasonality to establish a no-COVID baseline. Pandemic era deviations were modeled with Poisson state fixed effects regressions. Five week moving window t-tests flagged synchronous mortality spikes across states, and a two-way fixed effects event study estimated the elasticity of OD deaths to EIP related income shocks. JP Morgan Chase checking balance data validated the income–mortality link.

**Results:** From MarchL2020 to May □ 2023, OD deaths exceeded baseline by 67 □ 571 (24.4%). Sustained elevation was positively associated with income (β □ ≈ □ 0.96), fentanyl seizures, unemployment, and COVID-19 case rates, and inversely with methadone distribution. Three short lived spikes aligned with EIP disbursements, raising daily deaths by up to 85 above trend. A 10% rise in relative income increased OD mortality 11%; national checking balance surges and OD deaths were tightly correlated (r □ ≈ □ 0.90).

**Conclusions and Relevance:** Pandemic era overdose deaths comprise continuing exponential growth, a COVID-19 related sustained rise tied to social disruption, and EIP linked spikes.

Future relief payments should consider staggered disbursement and concurrent harm reduction measures to mitigate overdose risk.

## INTRODUCTION

Drug overdose (OD) deaths in the United States have followed a steadily increasing trajectory over the past four decades, characterized by exponential growth punctuated by intermittent periods of acceleration driven by changes in drug supply and use patterns.^1,2^ Prescription opioids, heroin, and most recently synthetic opioids such as fentanyl have each contributed to episodic surges in mortality.^3-5^

During the COVID-19 pandemic, OD deaths climbed sharply, accounting for a substantial portion of the overall decline in U.S. life expectancy.^3^ Multiple disruptions caused by the pandemic may have intensified vulnerability to drug overdoses, including heightened social isolation, unemployment, restricted access to healthcare, and changes in illicit drug markets.^5-7^ Some of these mechanisms disproportionately affected specific demographic groups and communities with limited healthcare resources, further magnifying preexisting health disparities. Concurrently, the availability of potent synthetic opioids such as fentanyl continued to expand, raising the risk of fatal OD across broader segments of the population.^2,8,9^

In addition, at the onset of the pandemic, major shifts in employment and income patterns occurred. The U.S. government introduced multiple policy interventions, notably three rounds of Economic Impact Payments (EIPs) between April 2020 and May 2021, intended to mitigate financial hardships resulting from COVID-19 shutdowns.^10,11^ Unlike unemployment insurance, which hinges on job loss, EIPs were distributed to a broad swath of the population regardless of employment status. Their amounts were often sizable relative to recipients’ baseline incomes, especially among low-income households.^10,12^ In prior research, large or lump-sum cash infusions have been linked to increased substance use, hospitalization, and overdose, which has been referred to as a “check effect”.^13-16^

Multiple factors may have contributed to the rise of OD deaths during COVID-19, however, there has been no detailed analysis of how the pandemic’s broader socioeconomic disruptions, the ongoing exponential increase in overdose mortality, and the timing and amounts of EIPs might have collectively influenced the surge in overdose deaths during COVID-19. This research fills that gap by using granular weekly overdose data and state-level data on multiple indicators, separating long-term overdose trends from shorter-term variability, thereby revealing that policy measures intended to alleviate economic distress may have inadvertently contributed to overdose risk.

## METHODS

Below we provide an overview of the data sources and methods implemented. For a detailed description, please refer to the Supplementary Materials.

### Data Sources

We use publicly available data from several sources to analyze factors associated with drug overdose deaths in the United States. Overdose deaths were defined using ICD-9 (1979–1998) and ICD-10 (1998–2023) codes for unintentional, suicide, homicide, and undetermined deaths.

Data on overdose mortality were obtained from CDC Wonder, CDC Rapid Release, and the National Vital Statistics System. State-level predictors included unemployment rates from the Bureau of Labor Statistics, personal income from the Bureau of Economic Analysis, fentanyl seizures from the National Forensic Laboratory Information System, methadone and buprenorphine distribution trends from the DEA’s ARCOS database, social distancing metrics from Google Mobility Reports, and COVID-19 cases and deaths from The New York Times. Information on the timing, numbers and amounts of payments by state for EIP rounds 1, 2, and 3 were reconstructed using reports from the Internal Revenue Service (IRS), the American Association of Retired Persons (AARP), and the Census Bureau Pulse Survey (Table S1). Checking account balance data were acquired from the JP Morgan Research Institute.

### Detrending OD deaths and adjusting for seasonal cycles (subcomponent 1)

To examine the patterns of excess mortality during COVID-19, we first projected the probable course of the national OD mortality curve without COVID-19 using a log-linear model with annual and monthly cycles, such that:

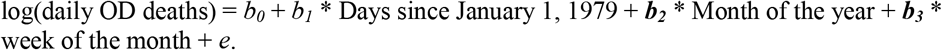

We included these terms because OD deaths are known to have an exponential trend and seasonal annual and monthly cycles.^1,17-19^ We fitted this regression model to OD deaths from January 1, 1979 until March 15, 2020, at which week COVID-19 Public Health Emergency (PHE) was declared and states began to implement shutdowns, projected it forward through December 30, 2023, and used this as a baseline to estimate excess OD mortality during COVID-19. We chose December 30, 2023, as the end-period for our analysis because the percent death records pending investigations increased sharply thereafter at the time of this analysis (Fig. S1).

### Association of OD deaths with state-level variables during COVID-19 (subcomponent 2)

To assess the association between socioeconomic and public health factors and OD deaths, we used Poisson generalized linear regression models with state fixed-effects to account for unobserved heterogeneity across states while examining time-varying predictors. The dependent variable was the monthly number of OD deaths, the main time-varying independent variables were percent population unemployed; percent time spent at home as a measure of social isolation; monthly income per person; methadone rate; buprenorphine rate; COVID-19 case and mortality rates; and fentanyl seizure rate. Standard errors were clustered at the state level and an offset term was added to adjust for population size, ensuring the results reflect per-capita effects.

### Periods of Synchronous Acceleration in OD Mortality across states (subcomponent 3)

To identify periods with significant synchronous acceleration in OD death rates across states, we performed a series of pairwise two-sided t-tests within a 5-week moving window. Each 5-week window was compared with the immediately preceding 5-week period, pairing observations by state, and Bonferroni-adjusted confidence intervals were used to correct for multiple testing. We slid this 5-week window forward one week at a time from January 1, 2018, through December 30, 2023, allowing the detection of short-term synchronous spikes in OD mortality across states.

### Association of EIP amounts and timings and OD deaths across states

To quantify the income elasticity of OD deaths—defined as the percent increase in OD mortality for each percent increase in income—we used the following two-way fixed effects (TWFE) model:

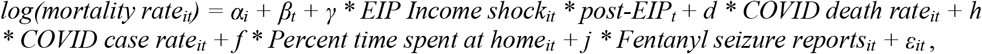

where *α*_*i*_ are state-level fixed effect for the *i* th state, *β*_*t*_ is weekly fixed effect for week *t, γ* represents the income elasticity of OD mortality. TWFE is a generalization of the standard difference-in-differences design that is used to control for both state-specific and time-specific confounders ^20-22^. To assess the parallel trend assumption and further explore how the amount of EIP payments influenced OD deaths across states, we employed a leads-and-lags (event-study) approach ^23^. We regressed weekly OD deaths on indicators for time periods before (leads) and after (lags) each EIP onset, using the week immediately preceding EIP distribution as the baseline.

### External Validation Using Checking Account Balances

As an external validation, we studied the temporal association between national weekly OD mortality (CDC data) and checking account balances from the JP Morgan Chase Institute, encompassing nearly 8 million families.^24^ Balances were stratified by age group (18–34, 35–54, 55+). We sought to determine if changes in checking account balances also coincided with increases in OD deaths, which would suggest that sudden infusions of disposable income, such as those occurring with EIP disbursements, may influence short-term OD mortality trends.

Furthermore, we estimated the relative risk of OD deaths by demographic characteristics (age category, sex, race, and urbanicity) following EIP initiation using Poisson regression models and interaction testing for subgroup-specific EIP effects.

## RESULTS

### Patterns of Excess OD Mortality During COVID-19

Fig. 1A reveals patterns of excess mortality during the COVID-19 pandemic. Between March 15, 2020 (the declaration of the COVID-19 public health emergency), and May 11, 2023 (its withdrawal), OD deaths remained elevated above the model-based projections (Fig. 1B). Notably, the CDC’s Vital Statistics Rapid Release provisional data, which uses a 12-month moving sum, obscured shorter-term monthly and annual variations (Fig. 1B). During this interval, three distinct and sudden transient spikes in OD deaths were observed, coinciding with the disbursement of the Economic Impact Payments (EIPs) (Fig. 1C). Supplementary Table S1 details the EIP schedules and amounts. Shortly after the conclusion of the public health emergency, OD deaths began to decline, approaching the curve predicted by our projection.

**Fig. 1.**
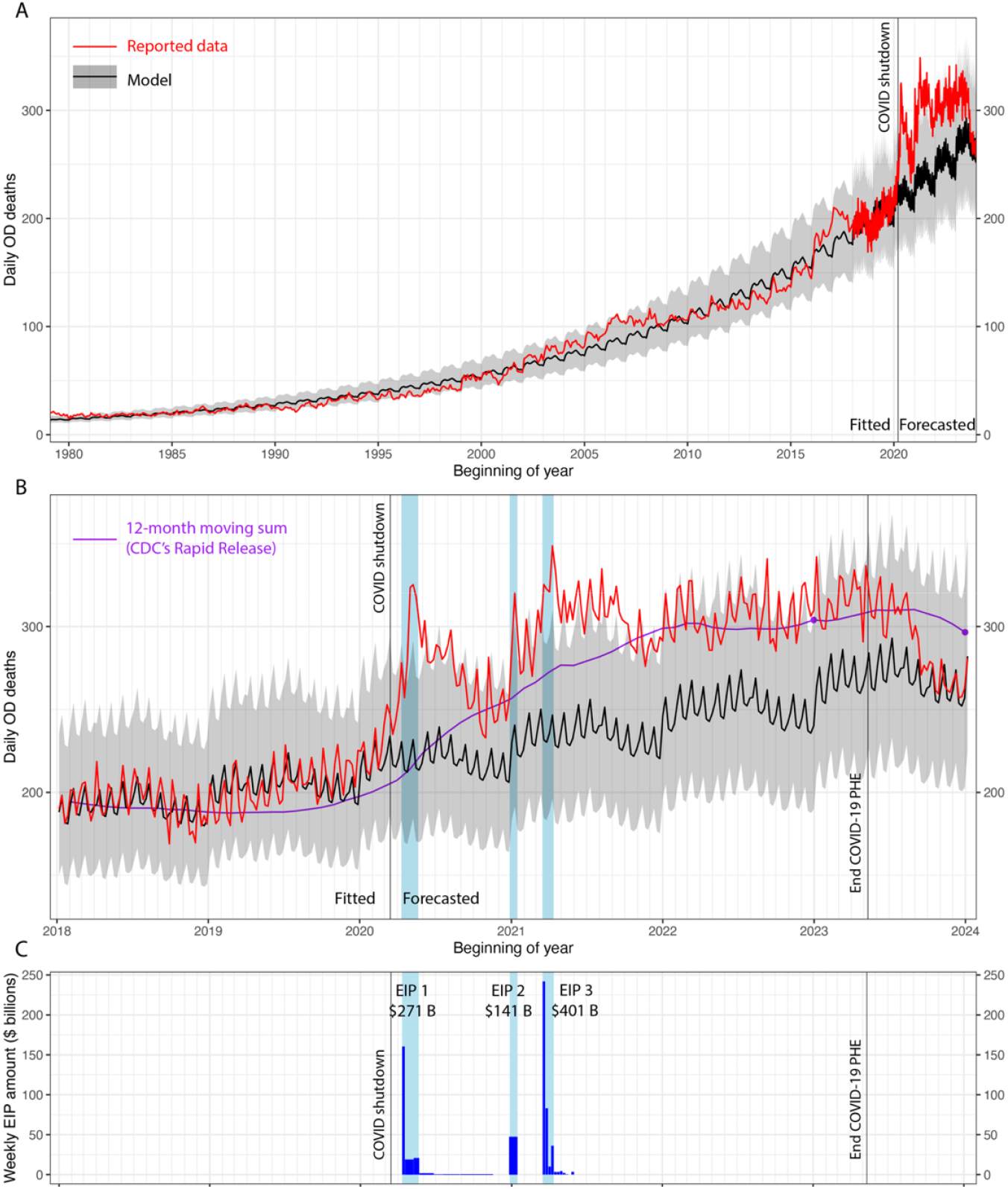
Monthly and weekly OD deaths reveal the three subcomponents of OD mortality during COVID-19. Panel A shows a model (black) fitted to reported OD deaths (red) from 1979 until March 15, 2020 (COVID-19 shutdown start), and then forecasted through December 30, 2023. The shaded gray region indicates the 95% predictive interval of the fitted model. Note that the darker lines in the far right of Panel A are due to the increased frequency of data points beginning in 2018 when weekly data became available. This period is shown in detail in Panel B. The purple curve shows CDC’s provisional counts which only report a 12-month moving sum. For example, the observation in January 2024 represents the total OD deaths for the 12-month period from February 2023 through January 2024 divided by 365 days. Panel C shows the weekly Economic Impact Payment (EIP) amounts in dark blue. The shaded light blue vertical ranges indicate the period in which 94% of the EIP payments were disbursed. The shaded light blue bars are extended upward into Panel B to facilitate comparisons. [PHE = Public Health Emergency].

The overall pattern of OD deaths during the pandemic can be decomposed into three distinct subcomponents: the long-term exponential growth pattern with superimposed annual and monthly cycles (subcomponent 1), a sustained rise followed by a decline in OD deaths during COVID-19 (subcomponent 2), and a series of transient spikes associated with the EIP disbursements (subcomponent 3).

### Subcomponent 1: Long-Term Exponential Growth with Annual and Monthly Cycles

This model fits the long-term OD death data with a high degree of accuracy (R^2^ > 0.98; Table S2). The model indicates that OD deaths increased at an exponential rate of 0.02% per day (p < 0.001). Due to this exponential component, daily OD deaths were projected to increase from 218 at the onset of the pandemic (March 15, 2020) to 270 by the end of the public health emergency (May 11, 2023). This reflects an average daily increase of +25 deaths over the period, attributable solely to the exponential trend.

Superimposed on this trend are recurring annual and monthly cycles. Annually, OD deaths peaked in March (+2.7%) and July (+2.9%) relative to January, with the lowest rates observed in December (-6.7%). Monthly cycles showed OD deaths peaking in the second week (+4.3%) and declining in the fourth week (-4.3%) relative to the first week of each month (Fig. 1A). These recurring patterns are consistent with prior trends observed before the pandemic.

### Subcomponent 2: Factors Associated with Sustained Increase During COVID-19

From March 2020 to May 2023, 344,428 OD deaths were recorded, compared to 276,857 deaths (95% CI: 274,589–279,126) predicted. This corresponds to an excess of 67,571 deaths (95% CI: 65,302–69,840), representing a 24.4% increase (95% CI: 23.4–25.4%) above expected levels.

Table 1 summarizes the factors associated with state OD death rates. Changes in income including wages, salaries, and government benefits such as EIPs—was the strongest predictor of OD deaths (β = 0.964, p < 0.001). Other significant predictors included fentanyl seizures (β = 0.204, p < 0.001), unemployment rates (β = 0.013, p < 0.001), and COVID-19 case rates (p < 0.001), while methadone distribution was negatively associated with OD deaths (β = -0.320, p = 0.035). Together, these predictors accounted for a significant portion of the variation in OD deaths across states, as evidenced by a McFadden’s Pseudo R^2^ of 0.34.

**Table 1.** Factors associated with increased in OD deaths across states.

| Variable | Coefficient | Standard Error | z value | p-value |
| --- | --- | --- | --- | --- |
| Year and month | 0.087 | 0.011 | 7.902 | < 0.001<br>*** |
| % unemployed | 0.013 | 0.004 | 3.735 | < 0.001<br>*** |
| % time spent at home | 0.004 | 0.002 | 1.822 | 0.068 . |
| Monthly income (detrended) | 0.964 | 0.110 | 8.798 | < 0.001<br>*** |
| Methadone rate | -0.320 | 0.152 | -2.108 | 0.035 * |
| Buprenorphine rate | 0.390 | 0.607 | 0.643 | 0.520 |
| COVID-19 case rate | 0.002 | 0.0004 | 5.733 | < 0.001<br>*** |
| COVID-19 mortality rate | -0.018 | 0.050 | -0.363 | 0.717 |
| Fentanyl seizure rate | 0.204 | 0.060 | 3.411 | < 0.001<br>*** |
| Observations | 3,600 |  |  |  |
| State fixed effects | Included |  |  |  |
| Offset | Log(population) |  |  |  |
| Log-Likelihood | -18,100.6 |  |  |  |
| Adjusted Pseudo R<sup>2</sup> | 0.932 |  |  |  |
| McFadden's Pseudo R<sup>2</sup> | 0.340 |  |  |  |
| BIC | 36,684.3 |  |  |  |

Model Statistics: Significance levels: \*\*\* p < 0.001, \*\* p < 0.01, \* p < 0.05, . p < 0.1

### Subcomponent 3: Transient Spikes During EIP Disbursements

During the three rounds of EIP disbursements, OD deaths exhibited short-term but pronounced spikes above the background rates defined by subcomponents 1 and 2. On average, subcomponent 3 contributed an additional +29 daily deaths during EIP weeks. These transient increases peaked during EIP 1 (+0.020 per 100,000), EIP 2 (+0.012 per 100,000), and EIP 3 (+0.014 per 100,000). Fig. 2A highlights the contribution of subcomponent 3, which coincided with EIP distribution periods and led to the highest levels of excess mortality above projections. The cumulative impact of these transient spikes was significant. For instance, during EIP disbursement periods, OD deaths reached a peak of +85 daily excess deaths above baseline.

**Fig. 2.**
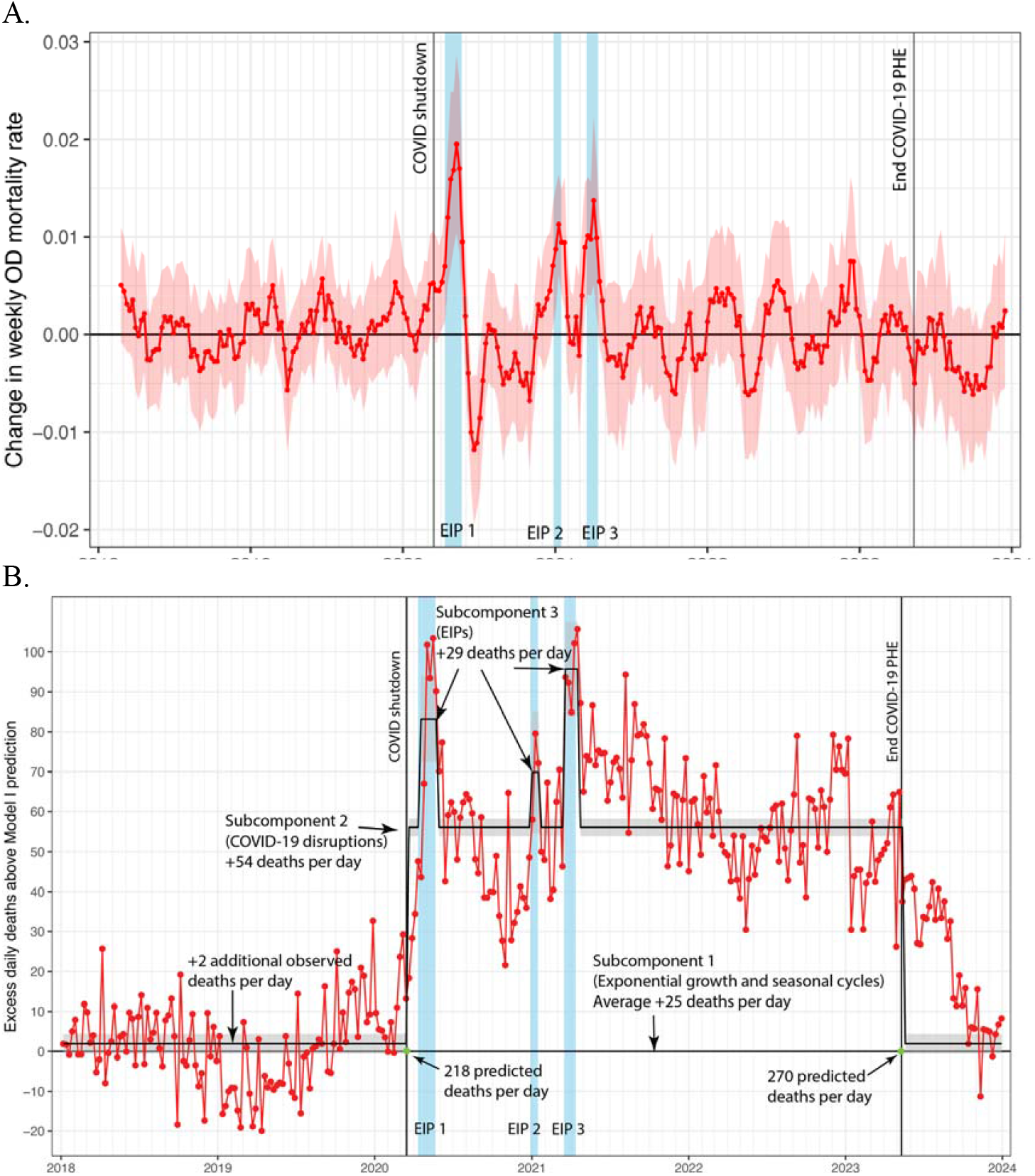
Temporal components of excess drug overdose (OD) mortality during the COVID-19 pandemic. **(A)** Week-to-week changes in OD mortality from 2018 to 2023 were gradual and asynchronous across U.S. states, except during periods when Economic Impact Payments (EIPs) were disbursed. The figure displays the mean weekly change and 95% confidence intervals in OD mortality rates across all 50 states. The light red area represents the 95% confidence interval, adjusted for multiple testing. Light blue vertical bars indicate periods during which 94% of EIPs were distributed. **(B)** Contributions of three subcomponents to the overall excess in OD deaths during the COVID-19 period. The red line shows excess daily deaths beyond the expected long-term exponential growth trend and seasonal cycles, modeled as Subcomponent 1 (baseline = zero). Subcomponent 2 captures the sustained elevation in mortality from the start of shutdowns on March 15, 2020, through the end of the Public Health Emergency on May 11, 2023. Subcomponent 3 is composed of three distinct spikes aligned with EIP disbursement periods.

Elevated OD deaths began to decline within weeks following each disbursement, returning toward the levels predicted by subcomponents 1 and 2. Notably, at the end of the public health emergency, daily OD mortality remained higher than pre-pandemic levels due to the continued influence of subcomponent 1, with 52 more daily deaths projected by the end of the observation period than at its onset.

#### Periods of Synchronous Acceleration During COVID-19 across states

Fig. 2B revealed significant week-over-week changes in OD death rates across states during the EIP disbursement periods. At the start of the COVID-19 shutdown, the baseline weekly OD mortality rate was 0.45 per 100,000. Early in the pandemic, OD mortality increased gradually by approximately 1% or 0.005 per 100,000 per week. During EIP disbursement periods, three distinct spikes in OD mortality were observed. The first spike (EIP 1, April-May 2020) reached an additional 0.020 per 100,000, while the second (EIP 2, January 2021) and third (EIP 3, March 2021) spikes reached 0.012 and 0.014 per 100,000, respectively. These represent weekly increases of approximately 4.4%, 2.7%, and 3.1% relative to the baseline weekly mortality rate. These accelerations indicate short-term but notable increases in OD deaths that were temporally aligned with the three EIP rounds. Outside these periods, OD death rates across states did not exhibit significant synchronous increases from 2018 to 2021.

#### Income Elasticity of EIP Disbursements on OD Deaths

The two-way fixed effects (TWFE) quantified the relationship between EIP income shocks and OD deaths. Income shock is calculated as the per capita EIP amount in a state relative to the median monthly household income in that state. The elasticity coefficient was 0.011 for the interaction between EIP income shock and post-EIP periods (P = 0.0004; R^2^=0.66) (Table S3). This suggests that for each 10% increase in relative income, OD deaths increased by 11%. To validate this elasticity estimate, we compared it against two external data sources: the JP Morgan checking account balance data and the Bureau of Economic Analysis (BEA) monthly income data. For each 10% increase in weekly checking account balances from JP Morgan’s data, OD deaths increased by 8% (95% CI=7.5–8.4%), and a 10% increase in monthly disposable personal income from the BEA was associated with a 23% (95% CI=20–27%) increase in OD deaths. Supplementary Fig. S2 reveals a lack of a systematic difference in pre-EIP trends, indicating no significant pre-trend in OD mortality prior to the income shock, but the lead coefficients were significantly higher than 0, indicating a sudden and persistent increase in overdose deaths in the weeks following the EIP disbursements. Furthermore, supplementary Fig. S3 shows that states with the greatest EIP income shocks experienced greater relative increases in OD deaths during COVID-19 compared to pre-COVID periods, with a correlation coefficient of r□= □0.51.

Additionally, several Western states formed a distinct cluster with lower EIP income shocks but higher OD mortality, likely due to higher fentanyl seizure rates, reflecting the spread of fentanyl to these regions during the pandemic.

#### Temporal Association Between Personal Bank Checking Account Balances and OD Deaths

Because both OD death subcomponents 2 and 3 were associated with personal income, we used checking account balances as an external source of validation. OD mortality and checking account balance data displayed tightly covarying temporal patterns across the three examined age groups (Fig. 3). Peaks in checking account balances in April/May 2020 and April 2021 were associated with corresponding spikes in OD deaths. Calculated with zero lag, the cross-correlations between the account balance and OD death time-series were rL=L0.901, 0.893, and 0.877 for age groups 18–34, 35–54, and 55+, respectively. Lagging the correlation of OD deaths to one week later than the checking balances improved the correlation to rL=L0.91 for the age group 35–54, but for the remaining comparisons, the correlation was highest at zero-week lag, signifying the temporal closeness between spikes in personal checking accounts and OD deaths. Notably, checking account balances and OD deaths began to increase in March 2020 with the start of COVID-19, reflecting a period of reduced spending and increased savings associated with the early pandemic.^24^ During this time, OD deaths increased in parallel. During EIP 1, and subsequently EIPs 2 and 3, both checking balances and deaths increased at a faster rate, especially for the 18–34 and 35–54 age groups. Figure S4 shows that relative checking account balances from JP Morgan began returning to their expected historical trend in 2023.

**Fig. 3.**
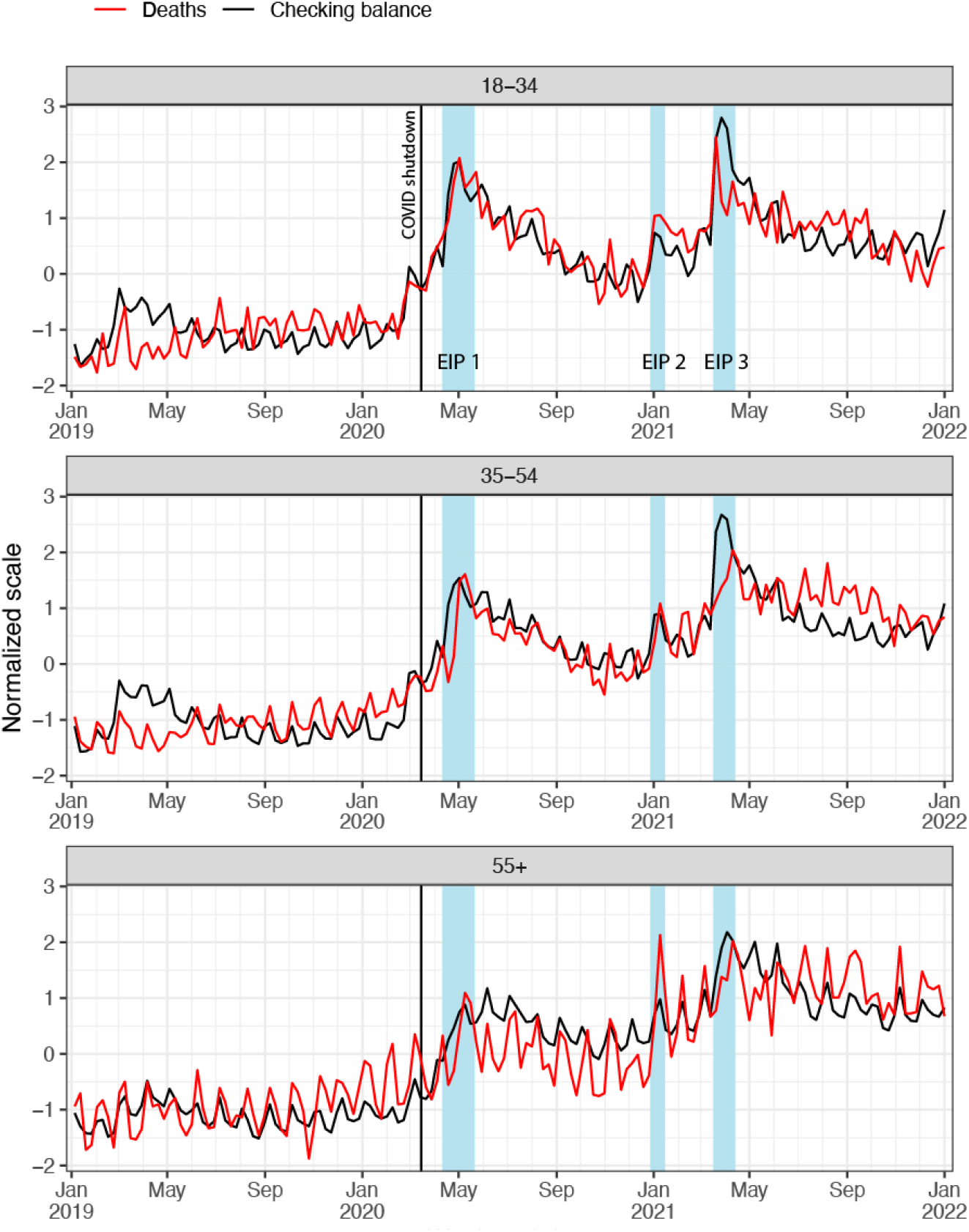
Temporal associations between drug overdose (OD) mortality and checking account balances during the COVID-19 pandemic. Parallel increases in drug OD deaths and checking account balances, both of which rose during the COVID-19 pandemic and transiently accelerated during periods of Economic Impact Payment (EIP) disbursement. Weekly OD death counts and checking account balances are shown for three age categories (18–34, 35–54, and 55+). Fluctuations in checking account balances occurred over several months and correspond to EIP intervals, marked by light blue shaded bars in which 94% of payments were distributed. Smaller monthly oscillations in checking balances may reflect routine income or social benefits. Data are normalized to enable comparison. [Fig. S4 reveals the return of checking account balances to historical trends in 2023.]

#### Relative Risk of OD Deaths by Demographics

Baseline risks were notably higher among individuals aged 35–44 (3.31 times the risk of those aged 15–24), males (2.30 times that of females), Blacks (1.23 times that of Whites), and those residing in metropolitan areas (1.13 times that of non-metropolitan areas). Post-EIP periods were associated with a roughly twofold increase in OD deaths (rate ratio = 1.98, 95% CI = 1.72–2.26), and the non-significant interaction terms (p > 0.05) indicate that this EIP-related increase was largely uniform across demographic subgroups (Table 2).

**Table 2.**
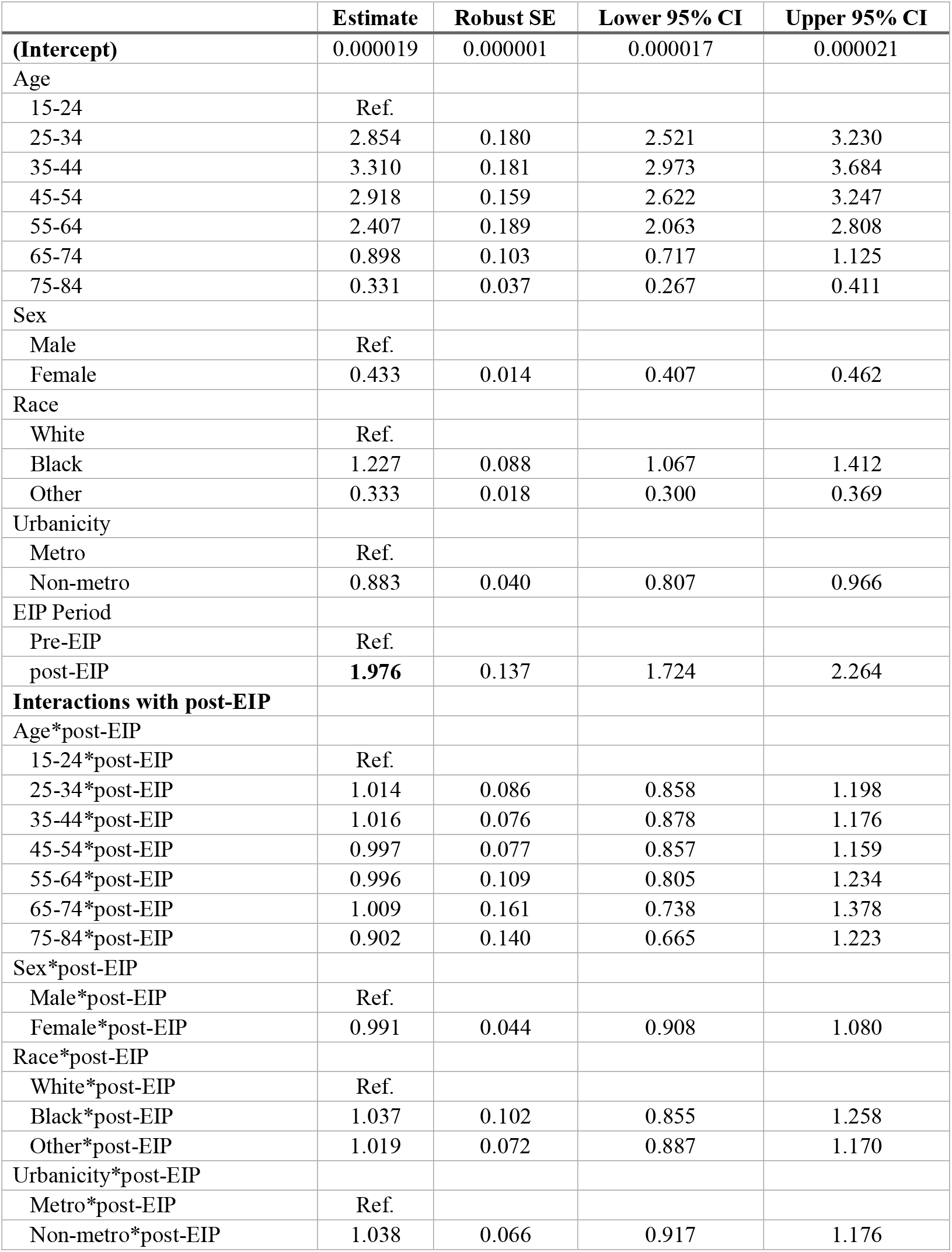
Post-EIP doubled baseline (pre-EIP) mortality rates across demographic groups. OD death rate ratios by demographics, EIP period, and their interactions are shown. Pre-EIP indicates the 5 weeks preceding each EIP and post-EIP indicates 5 weeks following each EIP.

## DISCUSSION

In this analysis, we decomposed national and state OD death curves into three subcomponents. We estimated excess deaths relative to a model representing the previously identified long-term exponential trend and the seasonal cycles.^1,17-19^ Following the exponential curve, OD deaths were expected to increase by +12% during COVID-19. However, the observed national OD death counts in this period far exceeded the expected counts, by an additional +24% beyond the expected +12%. We identified additional temporal patterns using weekly OD data that were not revealed from the 12-month moving sum of deaths of CDC’s Vital Statistics Rapid Release data (Fig 1B). While this 12-month format provides a valuable metric of trends over time, it obscures informative short-term changes. By plotting and analyzing the data as weekly counts (Figs. 1-4 and Table S4), we were able to identify three temporal subcomponents of the increase of OD deaths during COVID-19. We define subcomponent 1 (contributing +25 deaths per day) as the continuation of the long-term exponential growth, subcomponent 2 (contributing +54 deaths per day) as the abrupt and sustained increase in OD deaths throughout the COVID-19 pandemic, and subcomponent 3 (contributing an additional +14 to +40 deaths per day) as a series of three spikes representing periods of rapid and synchronous accelerations in OD deaths associated with the EIP disbursements. We specifically investigated the details of the timing and amount of EIP disbursements in relation to OD mortality.

### Subcomponent 1

The 40+ year steady exponential growth in OD death rates has been well described, but its causes have not yet been elucidated.^3,25^ A succession of different drugs predominated in this growth, including prescription opioids, then heroin, and most recently fentanyl (see Fig. 4 for a Conceptual Model). Note that the long-term exponential pattern in Fig. 1A has been temporarily disturbed by two previous transient upward deviations, one in 2006 driven by a transient increase in cocaine deaths ^1^ and a surge in 2016 and 2017 driven by carfentanil deaths ^26^. Probable drug-independent demand side explanations of this long-term growth involve societal structural factors such as poverty, income disparities, and despair ^27^. It is likely that supply side forces such as increased drug production and distribution efficiencies and profit are also involved. Whatever these long-term deep drivers of the exponential growth of OD deaths prove to be, we propose that they continued to operate uninterrupted during COVID-19 as subcomponent 1. The increase in fentanyl availability, which began at least four years before COVID-19, continued to be significantly associated with increased OD deaths during COVID-19.

### Subcomponent 2

The literature provides multiple hypothesized mechanisms through which COVID-19 may have increased OD deaths.^5-7^ While social isolation may have reduced some substance use among the youth,^28^ the negative impact of stay-at-home orders and social isolation are widely believed to have increased the level of stress and mental health impact especially among those who use drugs.^6,29,30^ In addition, job losses increased during the pandemic, which have been linked to decreased sense of purpose and increase OD deaths in Appalachia during the prescription opioid epidemic.^31-33^ COVID-19 has also disrupted delivery of health services and especially for drug treatment programs and changes in treatment supplies.^34-37^ While both OD and OD deaths increased in most states in the United States during COVID-19, diagnoses of substance use disorder declined, possibly indicating disruptions of access to health care.^38^ Furthermore, COVID-19 may have disproportionately increased the social, economic marginalization of some racial and ethnic minorities and may have restricted access to treatment and increased risk of OD deaths.^8,9^ Furthermore, the COVID-19 pandemic may have altered the supply and/or the quality of illicit drugs.^39^ In addition, we found several of these previously hypothesized factors to be significantly associated with increased OD deaths during COVID-19, including COVID-19 case rates, unemployment, increased income, and disruptions in opioid treatment access. We incorporated these measures of social disruption as contributing to subcomponent 2.

### Subcomponent 3

During the COVID-19 pandemic, the US government’s CARES Act provided EIPs and refundable tax credits totaling approximately $817 billion.^41^ While the EIPs offered critical financial relief during the pandemic, several features of these payments—especially their synchronous and rapid disbursement—may have inadvertently contributed to surges in OD deaths. A study using Ohio data shows a temporal correlation between the first EIP and increase in OD deaths in Ohio.^16^ The EIPs were based on income level, household composition, and number of children, without directly accounting for individuals’ actual loss of income or employment.^42,43^ As a result, median personal income increased by as much as 70% during the disbursement periods,^24^ a relative “positive income shock” that was greatest in low-income populations already at higher risk for substance use and OD.^27^

Large, lump-sum payments, including other pre-COVID programs such as stimulus checks and Supplemental Security Income, have historically been linked to increased risk of drug use, OD, hospitalizations, and deaths—often referred to as the “check effect.” In contrast, some studies have shown little or no association with OD deaths where payments were asynchronous and designed to replace actual lost income, as was the case with the Canada Emergency Response Benefit (CERB).^44^ Consequently, the synchronicity of EIP disbursements—wherein many recipients were paid in a narrow time window—may have exacerbated substance use–related harms, especially given the unprecedented amount and broad eligibility of these payments.^10,13^

**Fig. 4.**
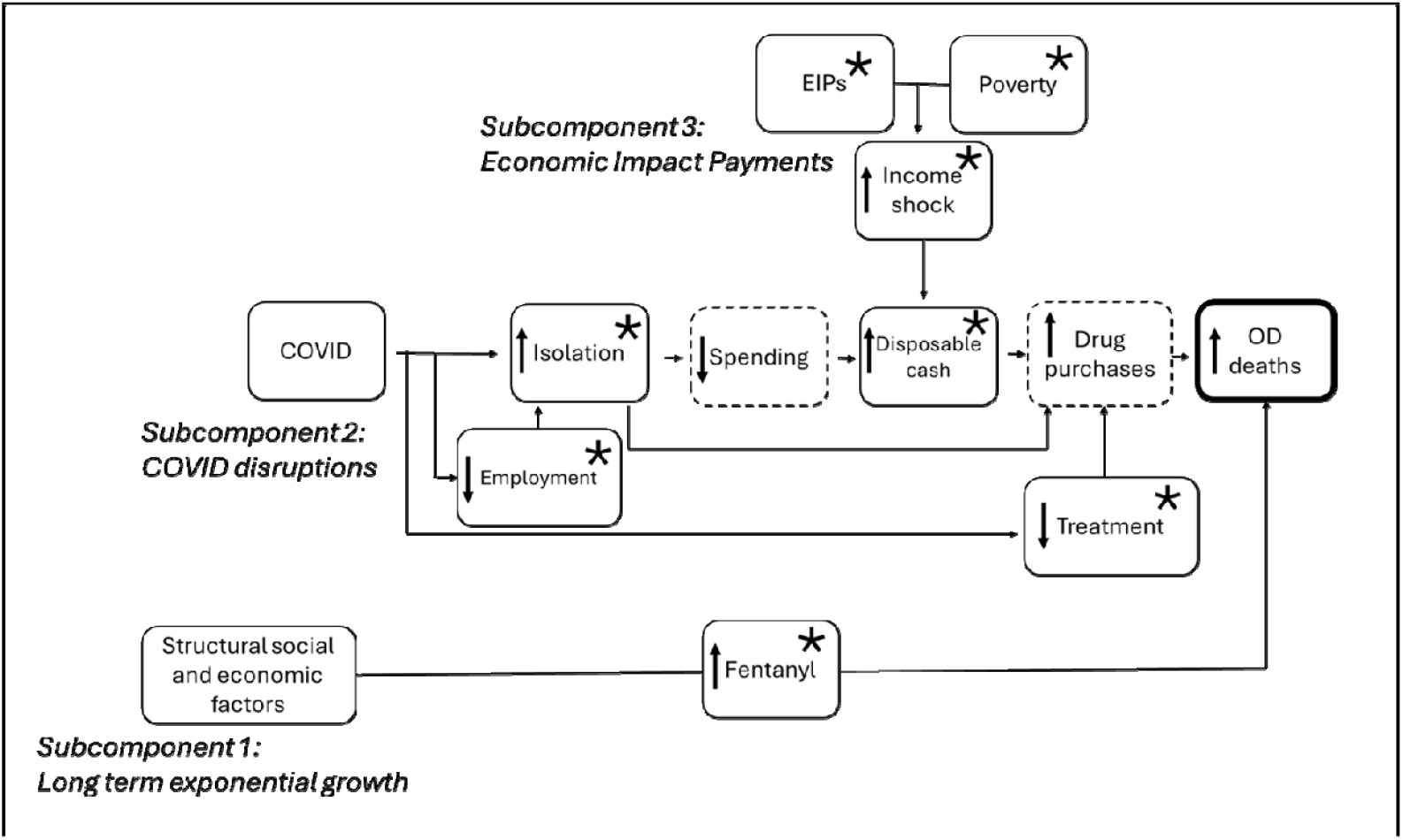
Conceptual model of the dynamics of increased OD deaths during COVID-19, showing the three subcomponents and their associated factors. Factors that are statistically associated with increased deaths in this study are marked with asterisks. Assumed factors are enclosed by dashed lines.

Income and OD deaths show complex dynamics. While unemployment insurance benefits, when calibrated to replace actual lost income, have been associated with lower overdose death rates,^33^ the EIPs were not contingent on job loss. Instead, the program delivered substantial funds rapidly to a wide swath of the population, possibly contributing to abrupt spikes in disposable income that can intensify OD risk, particularly within vulnerable groups. The timing and frequency of payments have repeatedly been identified as key factors influencing drug use and related harms,^13^ suggesting that alternative reimbursement regimes—such as smaller, more frequent, or more asynchronous payments—could help reduce the risk of overdose surges.

Despite the one harmful link between EIPs and OD deaths identified here, EIPs undoubtedly provided essential benefits to population health and well-being during the pandemic, such as helping households secure food and housing.^45,46^. Any evaluation of EIPs should carefully weigh all impacts—positive as well as negative—and consider whether future stimulus or relief efforts could be structured to mitigate OD risks. For instance, the fact that OD mortality accelerations were faster during EIP 2 and 3 than during EIP 1 could partly be due to expedited disbursements once the IRS logistics were refined ^47,48^ By EIP 2 and 3, an overwhelming majority of eligible individuals—particularly those below the Federal Poverty Line—received funds very quickly, whereas EIP 1 distribution faced initial delays for people without tax filing or direct-deposit information.^47,48^ More synchronous payments can mean large, one-time cash infusions arriving simultaneously, which may disrupt drug use patterns in high-risk communities.^49^

Additionally, although EIP-driven increases in disposable income are significant, other factors such as increased fentanyl availability and ongoing COVID-19–related disruptions also likely played major roles in the rise in OD deaths. Notably, checking account balances in many households remained elevated above pre-pandemic levels even outside EIP windows, reflecting broader shifts in spending and saving.^24^ These sustained changes in disposable income could have exacerbated or interacted with fentanyl prevalence to amplify OD deaths.

In May 2024, the CDC reported a 3% decline in OD deaths from 2022 to 2023.^50^ Though welcome, it remains unclear whether this decrease simply reflects a post-COVID return to the pre-pandemic OD death growth trends of subcomponent 1, or if it is an early indicator that overdose deaths might be dropping to levels below those forecasted before COVID. In this study, we used weekly overdose data, and integrated data from multiple sources to assess the association between overdose deaths and various state-level factors, including changes in COVID-19 incidence and mortality, unemployment, income, social isolation, amounts of methadone and buprenorphine sold, fentanyl supply, and the timing and amounts of economic impact payments (EIP). Understanding these factors may help evaluate the post-COVID OD death levels in the United States including the recent decline, and improve the assessment of future policies.

Certain limitations should be noted in our study. First, we lack a single source for fine resolution temporal OD death data in the United States especially towards the end of our observation period where death statistics are still provisional. We carefully combined three publicly available data sources from the CDC and verified the quality of these data as shown in the Supplementary Materials. Second, precise data on some of the factors associated with OD deaths were not available. For example, although we explicitly considered variations in the illicit drug supply over time, the available NFLIS data is limited because it records just the number of seizures reported by law enforcement agencies without providing any details on the actual amounts of the drugs seized. In addition, monthly, state-level data were not consistently available for all measures, and our reliance on fentanyl seizure data may underestimate actual supply. Third, by restricting our assessment of subcomponent 3’s impact to deaths occurring only within the EIP distribution windows, we likely underestimated their full effect. However, without a justifiable way to define the entire duration of the EIPs’ influences, we opted to use the distribution windows as a conservative estimate. Fourth, while the pronounced temporal alignment between EIP timing and OD death surges is suggestive, it does not establish definitive causality.

However, the rushed rollout of EIP 2—due to the impending expiration of certain CARES Act provisions—offers a quasi-experimental context wherein the timing was driven by external legislative deadlines rather than overdose trends.^51^ This feature reduces the likelihood of confounding factors influencing the observed association. Incorporating diverse data sources strengthened our findings, though the JP Morgan checking account information was limited by a lack of detailed geographic granularity. Finally, we limited our study to the United States. A cross-country comparison may further our understanding of the complex relation between OD deaths and factors associated with COVID-19.

In summary, this study identifies three distinct components contributing to the increase in drug overdose deaths in the United States during COVID-19. Applying this dynamic framework to analyze the fluctuations in overdose deaths offers valuable insights that can inform and enhance future drug overdose prevention and control policies. In addition, while this analysis highlights a harmful association between EIP disbursements and OD deaths, it should not overshadow the overall benefits of the EIPs in alleviating widespread economic hardship. Rather, our findings raise concerns about payment design features—especially the magnitude, frequency, and synchrony of disbursements—in shaping outcomes for high-risk populations. Future stimulus programs might benefit from incorporating deliberately asynchronous or gradual payment structures. Also, concurrent emergency drug use harm-reduction programs could be supported. These and other efforts should be implemented to minimize the risk of OD spikes without undermining the broader positive impact of emergency financial support.

## Data Availability

All data produced are available publicly from the indicated sources

